# Modeling the COVID-19 epidemic in Okinawa

**DOI:** 10.1101/2020.04.20.20071977

**Authors:** Simone Pigolotti, Davide Chiuchiu, Paula Villa Martin, Deepak Bhat

**Affiliations:** Biological Complexity Unit, Okinawa Institute of Science and Technology Graduate University, Onna, Okinawa 904-0495, Japan

## Abstract

We analyze current data on the COVID-19 spreading in Okinawa, Japan. We find that the initial spread is characterized by a doubling time of about 5 days. We implement a model to forecast the future spread under different scenarios. The model predicts that, if significant containment measures are not taken, a large fraction of the population will be infected with COVID-19, with the peak of the epidemic expected at the end of May and intensive care units having largely exceeded capacity. We analyzed scenarios implementing strong containment measures, similar to those imposed in Europe. The model predicts that an immediate implementation of strong containment measures (on the 19th of April) will significantly reduce the death count. We assess the negative consequences of these measures being implemented with a delay, or not being sufficiently stringent.

## I. INTRODUCTION

The first cases of COVID-19 epidemic in Okinawa were detected in mid-February 2020. After these few early cases, the epidemic was seemingly contained. The situation in Okinawa in February 2020 reflected a more general Japanese scenario where the epidemic was growing at a very slow pace, in contrast to the exponential growth in the case counts observed first in China, then in Europe, USA, and worldwide.

The situation in Japan and in Okinawa has drastically worsened since the end of March and beginning of April 2020. In this manuscript, we focus on the Okinawan case. Current data reveal that the number of cases in Okinawa in April 2020 has been growing exponentially, with a doubling time of about 5 days, see Fig. 1. This doubling time is very close to the one characterizing the early spreading dynamics in Wuhan, China [2, 3]. The last few data points seem to slightly depart from the exponential curve. It is difficult to assess at this stage whether this departure is due to early containment measures, or simply due to fluctuations. In the following, we shall first discuss a scenario in which no effective containment measures are in place. This scenario is idealized, as some measures are already in place and stricter measures will likely be implemented. However, it will serve us as a baseline to assess the effectiveness of containment measures. We will consequently consider scenarios where different containment measures are in place.

**FIG. 1:**
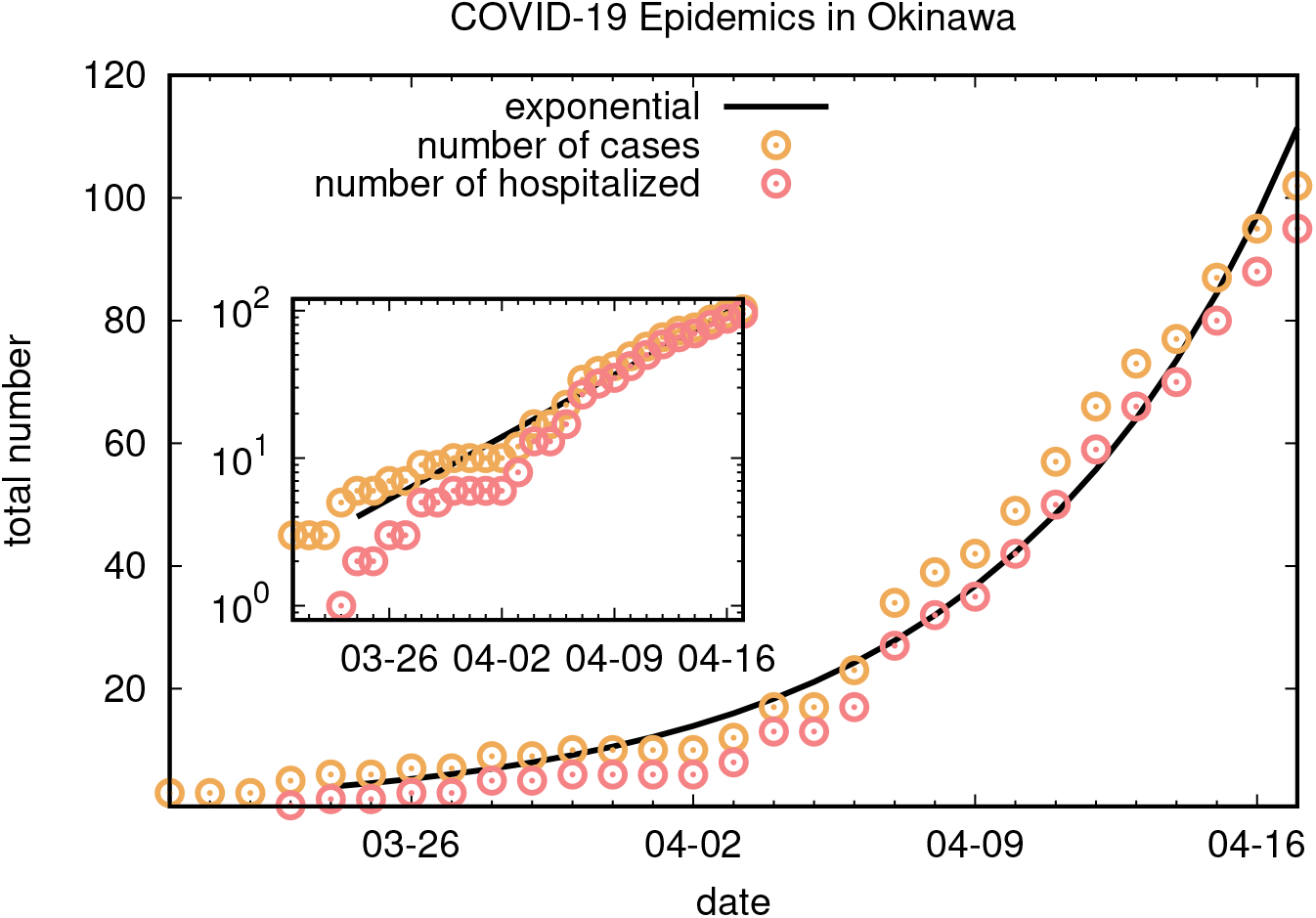
COVID-19 epidemic in Okinawa. The points are cumulative data from the Okinawa prefecture [1]. The black line is an exponential with doubling time 5 days, for comparison. The inset shows the same data in a linear-log plot.

## II. RESULTS

### A. Model - no containment

Our main tool is a model designed to predict and track the spread of the COVID-19 epidemic in a given region [4]. The model is a Susceptible-Exposed-Infected-Recovered (SEIR) model with realistic parameters for COVID-19 epidemics. Each of these categories is divided into age groups to account for the observed age-dependent risks [5]. The model permits to describe the import of cases from outside the region, assesses the case load on hospitals and intensive care units (ICUs) and includes a mortality risk due to ICUs exceeding capacity. It has recently been used to assess the epidemic risk in the Chicago, USA metropolitan area [6]. These characteristics make it an ideal model for our aims. We also developed an alternative SEIR model similar to that in [3] to verify the robustness of our results.

We adapted the model to the Okinawan scenario. To this aim, we used currently available estimates on COVID-19 parameters such as its incubation time and serial time [7–9], information about air travelers to Okinawa, data on age distribution of the Okinawan population, and daily number of confirmed and hospitalized cases in the Okinawan prefecture [1]. The model currently does not implement a regional age distribution, so that our simulations are performed using the Japanese age distribution. We shall discuss the potential impact of the different age distribution in Okinawa and Japan. Parameter choices and their rationale are further discussed in Methods.

We first run the model in a no-containment scenario, where the epidemic will keep spreading with the rate observed in Fig. 1 until herd immunity is reached. A crucial parameter to describe an epidemic is *R*_0_, the average number of infection caused by one infected person. We find that an *R*_0_ close to 2.5 provides a good fit of rate of increase in the number of hospitalized case as a function of time. Beside the parameters discussed in Methods, we had to make an assumption about the initial number of cases. We conventionally set the start of the simulation on the 15th of March 2020, a few days before the recent outbreak. We adjusted the initial number of cases so that the model fitted the number of hospitalized patients. Such procedure leads to estimate that in Okinawa there were about 10 cases on the 15th of March.

When fitting the number of hospitalized cases, the model predicts a staggering number of unseen cases. In particular, the model infers that by the 15th of April there were several thousands of infected and already recovered people in Okinawa – a number about 50 times larger than the number of confirmed cases. To understand this discrepancy, we compared with estimates from Europe. A recent study [8] performed a Bayesian analysis to estimate the percentage of infected populations in European countries on the 15th of March, 2020. We compared their results with the numbers of confirmed cases in Table III. This analysis reveals similar ratios between estimated and confirmed cases, especially in countries that did not perform extensive testing. In any case, a more conservative estimate of the number of infected cases would not significantly alter our results. In the absence of containment measures, the model predicts that a large fraction of the Okinawan population will bereached by the contagion, with a peak occurring roughly at the end of May. At the peak, the model predicts about 5,000 critical patients requiring an ICU bed, far more than the existing capacity. The predicted cumulative death count by the end of the epidemic exceeds 20,000 people. A large fraction of these deaths are predicted to occur after the peak in the number of infections. A crucial assumption underlying this number is that the death rate of critical patients is doubled if no ICU bed is available. This means that about half of the deaths predicted by the model are ascribed to ICUs exceeding capacity.

The alternative model we implemented predicts a number of deaths exceeding 10,000, with a similar peak time. The alternative model does not include the additional risk due to ICU overflow, so that the predictions of the two models are consistent. This supports that our result is robust with respect to model idiosyncrasies. We used the alternative model to estimate the error made in using the Japanese age distribution as a proxy of the Okinawan one. The model predicts that this approximation leads to an overestimate of the death count by about 20%. This overestimate should be taken into account also in the scenarios discussed in the next section.

One choice that could affect the result is fitting the curve of hospitalized patients, as this makes the results dependent on the threshold for hospitalization in Okinawa. However, changing this threshold would shift the result by a few days to one week, without affecting our main conclusion. We also remark that a death toll of 10000 plus consequences of ICU capacity is consistent with an order of magnitude estimate, as the average mortality rate of COVID-19 is estimated to exceed 1% [10]. In case of a continued, uncontained spreading, the contagion will affect a large fraction of the Okinawan population, which consists of about 1.4 million individuals.

## III. CONTAINMENT SCENARIOS

The scenario of the previous section does not take into account containment measures set up in response to the epidemic spreading. Some measures have already been taken, including suspension of some public events [11] and closing of schools [12]. We summarize the effect of containment by the number *R*_*t*_ of infections caused by each infected individuals in the presence of these measures. Due to containment, *R*_*t*_ is expected to be is reduced compared to *R*_0_, which is the same parameter in the absence of containment measures. The percentage reduction quantifies the combined effectiveness of containment measurements. In this section, we consider scenarios with different values of this reduction factor, and different implementation times. In the Discussion section, we then assess which concrete measures might lead to these reduction factors, based on the experience in European countries.

We first ran the model in a hypothetical scenario where containment measures result in a *R*_*t*_ being 65% smaller compared to *R*_0_. This percentage is chosen so that *R*_*t*_ is below one, meaning that the measure is sufficient to stop the epidemic.

The model estimates that such containment measures, if implemented on the 19th of April, would bring down the death count to about 1000, including the effect of ICU overload, see Fig. 3, left. In this scenario, the ICU capacity required at the peak is of roughly 170 beds. To assess the effect of a delay in implementing containment, we simulated an alternative scenario where the same containment measures are implemented on the 1st of May, see Fig. 3, right. In this case, the model predicts about 4000 deaths, with a required ICU capacity at the peak of about 800 beds.

**FIG. 2:**
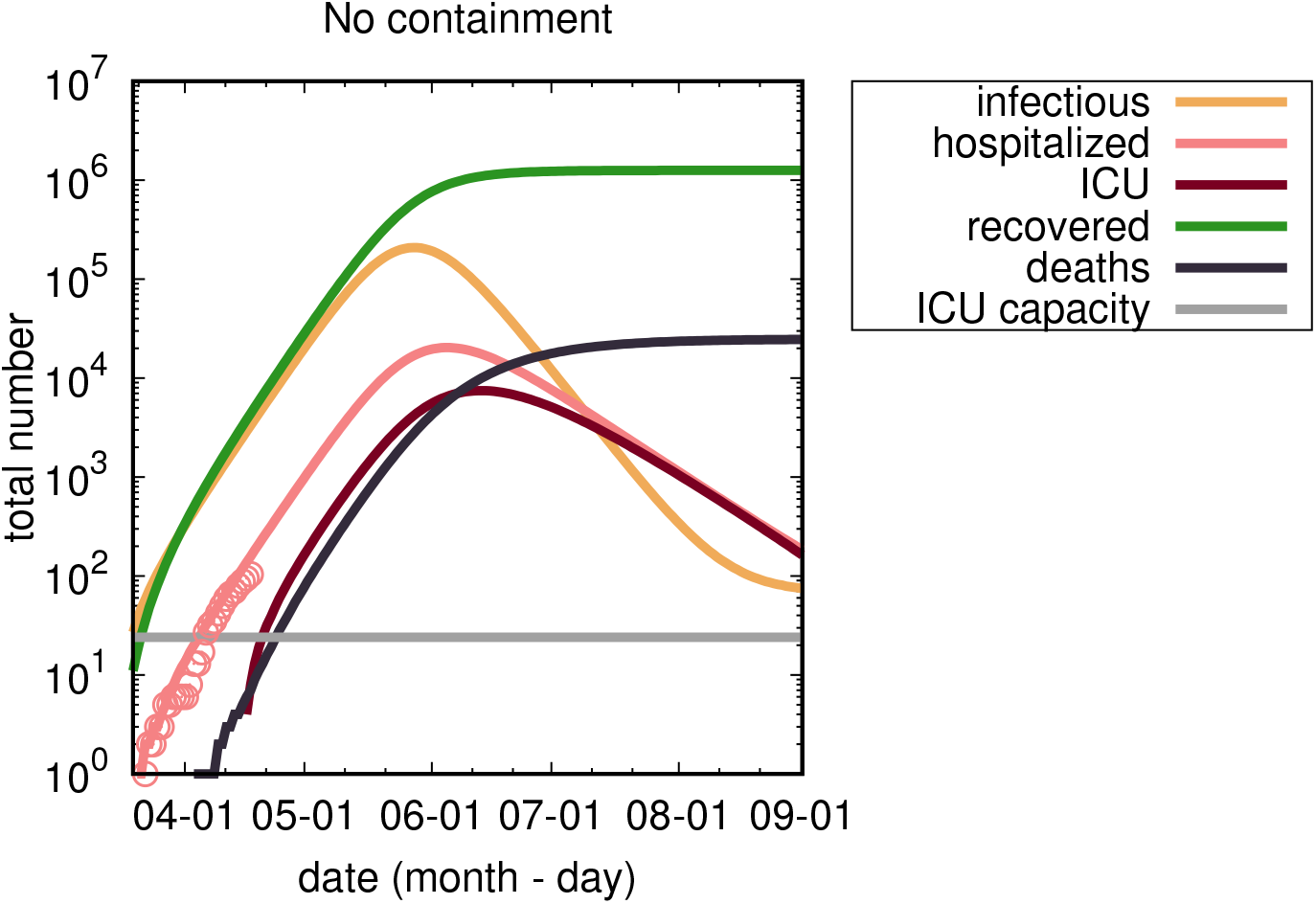
Model results for the no containment scenario. y-axis is logarithmic. Points are the number of hospitalized infected patients in Okinawa as reported in [1]. Lines are modeling predictions (color scheme explained in the legend). The gray horizontal line denotes the current ICU capacity in Okinawa (24 beds). The dark red line (ICU) denotes the number of critical patients requiring ICU beds.

**FIG. 3:**
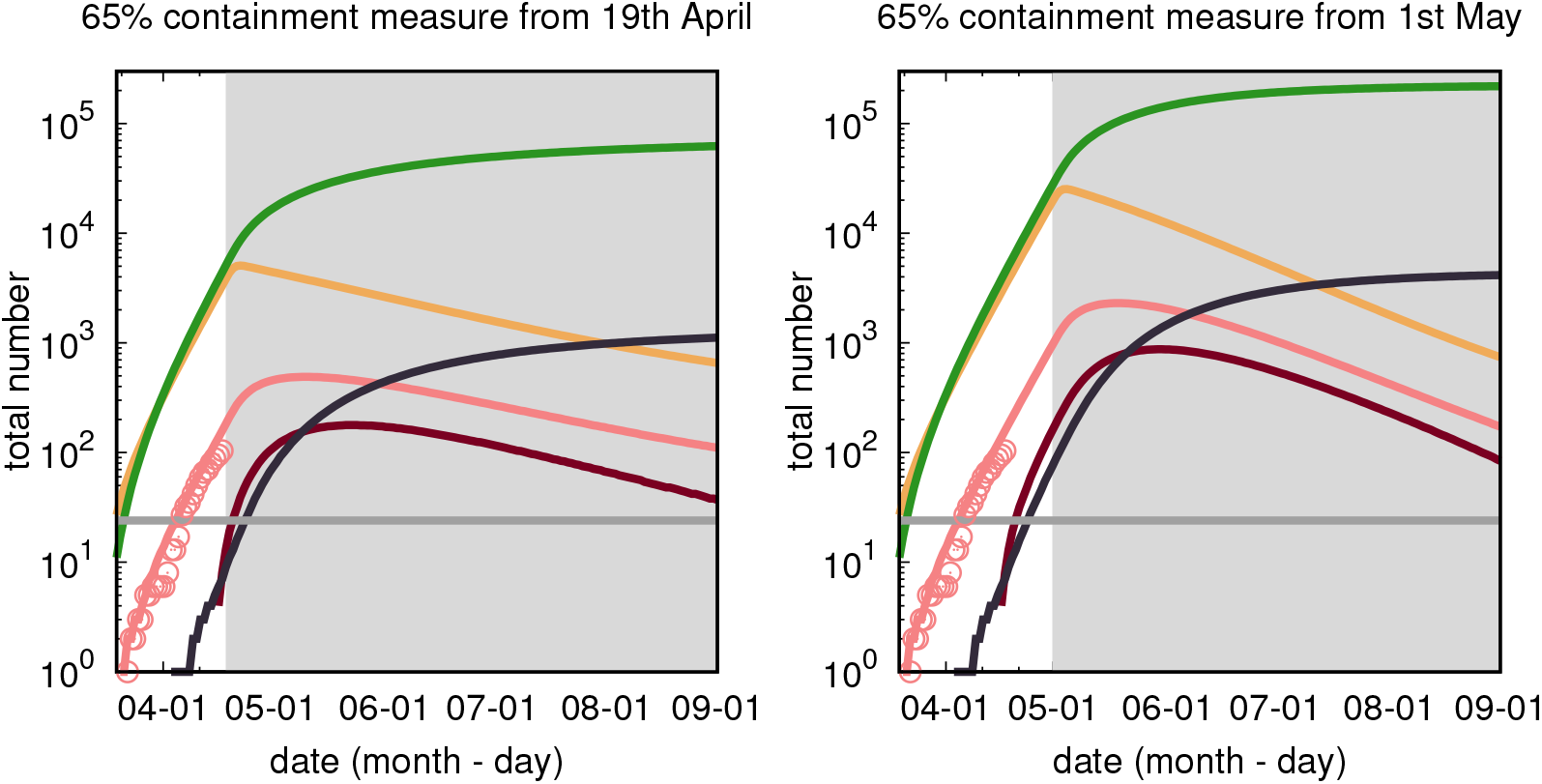
Model results for containment measures reducing *R*_*t*_ by 65% with respect to *R*_0_. Left panel: measures enacted from the 19th of April, 2020. Right panel: measures enacted from the 1st of May, 2020. Different curves are different model predictions as in Fig. 2. The gray region marks the time interval where containment measures are imposed.

In the containment scenarios discussed so far, it is crucial that the implemented measures reduce *R*_*t*_ below one. To show that this is the case, we simulated a scenario in which containment measures implemented on the 19th of April reduces *R*_*t*_ by only 40% with respect to *R*_0_, see Fig. 4. Our model predicts that such measures will significantly slow down the epidemic, delaying its peak to the beginning of July. However, in the presence of such measures the model predicts a total death count at the end of the epidemic of about 14,000 and a required ICU capacity at the peak of about 2700 beds.

**FIG. 4:**
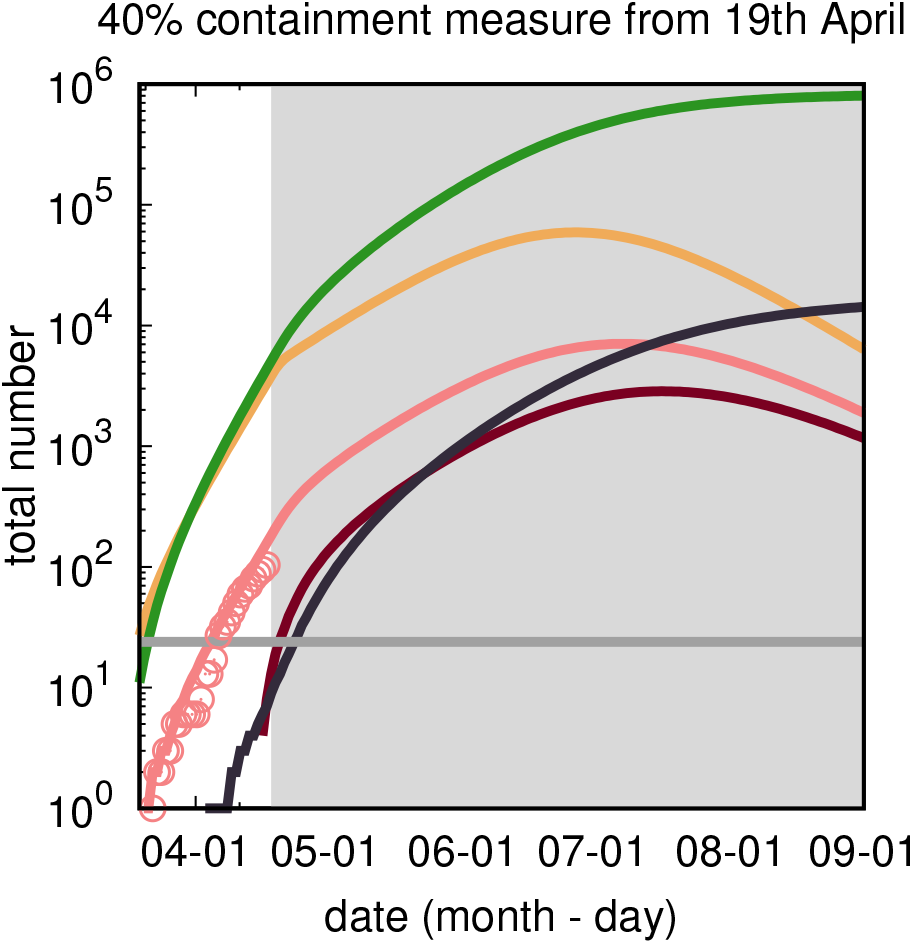
Model results for a containment measure reducing *R*_*t*_ by 40% with respect to *R*_0_. The measure acts since the 19th of April, 2020. Different curves are different model predictions as in Fig. 2. The gray region marks the time interval where containment measures are in place.

## IV. DISCUSSION

Our analysis of the spread of the COVID-19 epidemic in Okinawa reveals that its initial exponential growth rate is similar to that observed first in Wuhan, and more recently in many European countries. A model based on information gathered from these cases predicts that a rapid and strong reduction in the rate of spread is necessary to avert scenarios in which the Okinawan health care system will not be able to cope with the epidemic.

Some measures to contain the epidemic have already been taken, partially following the declaration of a state of emergency over the COVID-19 epidemic by the central government on the 8th of April 2020. Measures taken in Okinawa include suspension of some public events [11], closing of schools [12], and increased controls at the airport. The mild reduction in the growth rate of the epidemic observed in the last days (Fig.1) could be due to these measures, if we take into account an expected lag of more than one week between the introduction of a measure and its measurable effects.

It is crucial to assess whether the measures in place are likely to have brought *R*_*t*_ below one. An analysis of containment measures introduced in different European countries attempts at dissecting the impact on *R*_*t*_ by these measures [8], see Table I. The median effect on *R*_*t*_ of measures such as school closure, increased social distancing and public events is rather moderate according to this analysis. It should be noted that these estimates are affected by a large variability, caused by data quality and different ways in which these measures have been implemented in different countries. A “lockdown” requires people to remain at home wherever possible, leaving their house for essential trips only (to buy food, medicine, and take daily exercise), and avoiding groups. The analysis in Table I shows that lockdowns in Europe have reduced *R*_*t*_ much more effectively than other measures. Combined with our modeling results, this leads to the conclusion that Okinawa should consider imposing soon some sort of lockdown.

**TABLE I:**
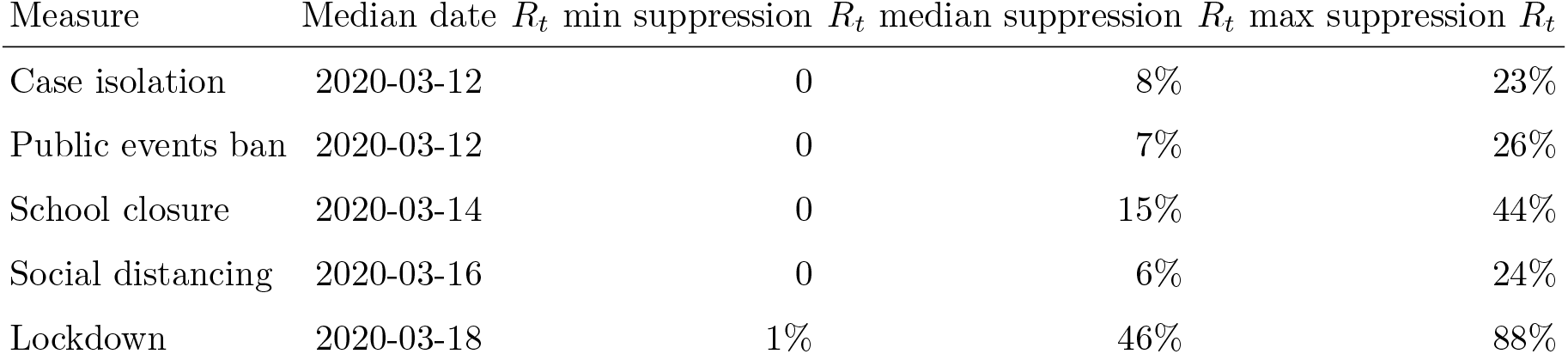
Estimated reduction in *R*_*t*_ from different suppression measures implemented in European countries. The reported range is a 95% confidence interval. Adapted from Fig.4 in [8].

Available data and the experience of European countries suggest that the containment scenarios predicted by the model are realistic and therefore more stringent measures should be considered in Okinawa. Nevertheless, the model we implemented are necessarily simplified and it will be possible to perform more accurate estimates once more information is available, for example about the potential seasonality of the epidemic. As the epidemic develops, we expect stronger measures to be taken, that will possibly skew the numbers away from the example scenarios we considered. It will be therefore important to update our analysis at these later stages, thereby assessing the efficacy of such measures.

## Data Availability

The manuscript includes data analysis and simulation results. Data are publicly available. Code for the alternative SEIR model can be made available upon reasonable request.

## Acknowledgments

This manuscript is a working document, not summitted for peer-review. It is distributed as an early version to provide a preliminary analysis during the epidemic spread. SP acknowledges Luca Ferretti for useful discussions.

## V. METHODS

### Parameters

- **Incubation time, infectivity period, and serial interval** The incubation time of COVID-19 is estimated to be around 5 days [7, 8]. During this period, pre-symptomatic transmission may occur, which is not included in the SEIR model. Recent analysis suggest a serial interval, i.e. a time between consecutive infection in a chain of transmission, on the order of 5 to 6 days [9]. To account for presymptomatic transmission and predict a correct serial interval, we fix an effective incubation time of 3 days and an infectivity period of 5 days.
- **R**_0_. Studies of the early Wuhan epidemics estimate *R*_0_ *≈* 2.2 [13], *R*_0_ *≈* 2.35 [7], *R*_0_ *≈* 2.57 [14]. More recent estimates tend to the lower end of this spectrum [3] and a slightly longer generation time. In order to reproduce the observed doubling time in the number of cases we determine *R*_0_ = 2.5. A more precise estimate of *R*_0_ from the doubling time would require more detailed information about the distribution of infection intervals [15].
- **Imported cases per day**. We estimate roughly 1000 passengers per day arriving at Naha airport in March and early April. The majority of this passengers are from mainland Japan. In Japan, during this period, there were on the order of 1000 confirmed cases. Including a correcion for a large number of unobserved case, similar to that in Table III, we assume that 1% of the Japanese population was infected during this time window. This assumption leads to about 10 infected passengers arriving at Naha airport per day. A more precise estimate would be important to describe more accurately the initial stage of the epidemic, but would not significantly affect its development once a large number of cases are present in Okinawa.
- **Population structure**. Population structure is an important determinant of COVID-19 severity. The model accounts for the official age distribution of the population, but currently only at a national level, i.e. it assumes that the age distribution in Okinawa is the same as in Japan. Okinawa has a younger age distribution than Japan, see Table. II. A comparison using our alternative SEIR model reveals that using the Japanese population structure instead of the Okinawan one leads to an overestimate of the death count on the order of 20%.

**TABLE II:** Comparison of the age structure in Okinawa with the entire Japan, from 2015 census.
- **ICU**

| Area | 0-10 | 10-20 | 20-30 | 30-40 | 40-50 | 50-60 | 60-70 | 70-80 | 80-90 | 90-100 |
| --- | --- | --- | --- | --- | --- | --- | --- | --- | --- | --- |
| Okinawa | 11.5% | 11.4% | 10.5% | 13% | 13.8% | 12.5% | 12.3% | 7.8% | 4.9% | 1.1% |
| Japan | 8.1% | 9.1% | 9.3% | 12.2% | 14.5% | 12.1% | 14.2% | 11% | 6.3% | 1.3% |

The number of ICU beds in designated medical institution in Okinawa is 24 according to [16].

## VI. RATIO BETWEEN OBSERVED AND REAL CASES

We report in Table III an estiamte of the ratio between observed and estimated number of cases in different European countries from [8].

**TABLE III:** The number of confirmed cases until 28th March versus inferred percentage of cases in the total population predicted in Ref.[8]) in European countries. The last column report the ratio between observed and inferred cases.

| Country | Confirmed | Inferred cases (% of total population) | $\frac{\text{Inferred}}{\text{Confirmed}}$ |
| --- | --- | --- | --- |
| Austria | 8271 | 1.1 | 11.96 |
| Belgium | 9134 | 3.7 | 46.90 |
| Denmark | 2201 | 1.1 | 28.93 |
| France | 37575 | 3 | 52.09 |
| Germany | 57695 | 0.72 | 10.45 |
| Italy | 92472 | 9.8 | 64.10 |
| Norway | 4015 | 0.41 | 5.53 |
| Spain | 73235 | 15 | 95.76 |
| Sweden | 3447 | 3.1 | 90.71 |
| Switzerland | 14076 | 3.2 | 19.65 |
| United Kingdom | 17089 | 2.7 | 107.14 |

## Notes

### Competing Interest Statement

The authors have declared no competing interest.

### Funding Statement

none to report.

